# The Adequacy of Vancomycin Initial Dosing in CRBSI for Hemodialysis Patients at Hospital Pakar Sultanah Fatimah, Muar, Johor

**DOI:** 10.1101/2025.03.13.25323881

**Authors:** Rohidayah Abd Majid, Er Pei San, Nurul Fazleen Abdul Malik, Mohd Afif Syazani bin Sulaiman, Nur Fatin Afiqah Fakhrul Anwar, Muhammed Luqmanulhakim Abu Bakar

## Abstract

**Background:** Catheter-related bloodstream infections (CRBSIs) are a common complication in hemodialysis (HD) patients with central venous catheters (CVCs), often caused by Methicillin-Resistant *Staphylococcus aureus* (MRSA). Vancomycin is the preferred first-line treatment for MRSA-related CRBSI. At Hospital Pakar Sultanah Fatimah (HPSF), the current protocol for CRBSI in HD patients involves a weight-based intravenous (IV) vancomycin loading dose (LD) of 25 mg/kg, followed by therapeutic drug monitoring (TDM) within 48 hours post-administration. This study evaluates the adequacy of the initial dosing regimen in achieving the target serum vancomycin concentration (SVC) of 15–20 mcg/ml.

**Methods:** A retrospective cross-sectional study was conducted among hospitalized HD patients receiving IV vancomycin from July 1, 2022, to July 31, 2023. Data were collected from TDM request orders in the Pharmacy Information System (PhIS). The primary outcomes analyzed were the proportion of patients achieving the target SVC and factors influencing vancomycin pharmacokinetics. Multiple linear regression was used to assess associations between SVC and variables such as time-to-first-sampling (TTFS), age, and initial vancomycin dose.

**Results:** A total of 106 TDM samples were analyzed. The mean vancomycin LD was 1568.2 mg (SD 303.6), with a mean TTFS of 41.2 hours (SD 11.8). Only 32.1% of patients achieved the target SVC, while 56.6% had subtherapeutic levels and 11.3% had supratherapeutic levels. Multiple linear regression identified TTFS (p=0.003), age (p=0.002), and initial dose (p=0.004) as significant predictors of SVC.

**Conclusion:** The current vancomycin dosing protocol at HPSF does not consistently achieve therapeutic SVC in HD patients with CRBSI. Increasing the LD to 25–35 mg/kg and optimizing TTFS within 24–48 hours may improve target attainment. Further studies are needed to validate these findings and refine vancomycin dosing strategies in HD patients.

## Introduction

End-stage renal disease (ESRD) is a significant global public health concern. In Malaysia, hemodialysis (HD) is the preferred treatment modality for ESRD. The prevalence of ESRD patients undergoing dialysis has increased dramatically, from 7,837 in 2001 to nearly 23,000 in 2010 [2]. By 2014, the total number of patients requiring dialysis in Malaysia had risen to 34,767, with an annual mortality rate of 11.6% among hemodialysis patients [3].

One of the major complications in hemodialysis patients with central venous catheters (CVCs) is catheter-related bloodstream infections (CRBSIs) [4]. It is estimated that 80% of incident hemodialysis patients rely on CVCs as their primary vascular access, with a significant proportion initiating dialysis with non-tunneled CVCs, particularly in developing countries [5].

Among the most commonly isolated pathogens in CRBSI cases are staphylococcal species, with Methicillin-Resistant Staphylococcus aureus (MRSA) emerging as a prevalent concern [6]. Vancomycin is frequently used as the first-line treatment for MRSA-related CRBSI [7]. However, due to limited data and the lack of standardized guidelines for vancomycin dosing and therapeutic drug monitoring (TDM) in hemodialysis patients, there is significant variability in vancomycin dosing regimens across healthcare institutions [8].

The mechanism of hemodialysis involves diffusion and ultrafiltration, which facilitate the movement of solutes across a semi-permeable membrane [9]. Dialyzers are categorized into two major types: high-flux and low-flux. Low-flux dialyzers, characterized by an ultrafiltration rate of <15 mL/mmHg/h, are primarily effective in removing small solutes through diffusion but have minimal clearance of middle-sized solutes, which are considered more toxic and difficult to eliminate by diffusion alone [10]. At Hospital Pakar Sultanah Fatimah (HPSF), hemodialysis procedures are conducted using low-flux dialyzers.

Currently, HPSF follows a weight-based vancomycin dosing protocol for CRBSI treatment in hemodialysis patients. This protocol consists of an intravenous (IV) loading dose of 25 mg/kg, followed by TDM within 48 hours post-administration to assess drug levels. To optimize clinical outcomes, it is crucial to evaluate the effectiveness of this protocol. Therefore, this study aims to assess the adequacy of the initial vancomycin dosing and the time-to-first-sampling (TTFS) in achieving the target serum vancomycin concentration (SVC) of 15-20 mcg/mL.

## Methods

This cross-sectional retrospective study was conducted among hemodialysis (HD) patients at Hospital Pakar Sultanah Fatimah (HPSF), Muar, from July 1, 2022, to July 31, 2023. The study included hospitalized end-stage renal disease (ESRD) patients aged 18 years and above who were undergoing hemodialysis and received intravenous vancomycin during admission as either empirical or definitive treatment for an infection.

A universal sampling method was applied, where eligible patients were identified through vancomycin Therapeutic Drug Monitoring (TDM) request orders in the Pharmacy Information System (PhIS). The medical records of these patients were reviewed to collect socio-demographic data, comorbidities, dialysis prescriptions, vancomycin dosing, and administration records. Additionally, time to first sampling (TTFS) and serum vancomycin concentration (SVC) following the initial vancomycin dose were retrieved from the TDM order in PhIS.

In this study, TTFS was defined as the TDM sampling time occurring 36 to 48 hours after the vancomycin loading dose, while a TDM request was defined as a TDM test order placed by a prescriber via PhIS.

Data analysis was performed using IBM SPSS Version 26. The weight-based initial vancomycin loading dose, TTFS patterns, and resulting SVC were analyzed descriptively. Categorical data were presented as frequencies (n) and percentages (%), whereas continuous data were reported as mean and standard deviation (SD).

To determine factors influencing SVC after the initial vancomycin dose at first TDM sampling, multiple linear regression analysis was conducted. Initially, simple linear regression was performed for all variables, and those with p < 0.25 were subsequently included in the multiple linear regression model. The regression results were expressed as regression coefficients (β) with 95% confidence intervals (CI), and statistical significance was set at p < 0.05.

This study was approved by the Medical Research and Ethics Committee (MREC), Ministry of Health Malaysia, with registration number NMRR ID-24-02412-PBR. Patient privacy and confidentiality were strictly maintained throughout data collection, analysis, and publication by ensuring no patient identifiers were included.

## Results

A total of 106 TDM samples were included in this study. The demographic and clinical characteristics of the patients are presented in Table 1. The mean age of patients whose TDM samples were analyzed was 56.3 years (SD 12.7). The distribution of male (52.8%) and female (47.2%) patients was nearly equal. The majority of patients were younger than 60 years old and of Malay ethnicity (85.8%).Most patients received vancomycin as a definitive treatment (93.4%), with only 6.6% receiving it empirically.

**Table 1:** Patient Demographic and Clinical Characteristics (n = 106 TDM samples)

| Characteristics | Value |
| --- | --- |
| Age,year,mean (SD) | 56.3 (12.7) |
| Age,year, n (%) |  |
| ≥ 60 | 46 (43.4) |
| <60 | 60 (56.6) |
| Weight, kg, mean (SD) | 67.4 (14.6) |
| Gender, n (%) |  |
| Male | 56(52.8) |
| Female | 50(47.2) |
| Race, n (%) |  |
| Malay | 91(85.8) |
| Chinese | 13(12.3) |
| Indian | 2(1.9) |
| Indication of vancomycin, n (%) |  |
| Empirical | 7 (6.6) |
| Definitive | 99 (93.4) |

Initial Dose of Vancomycin and Serum Vancomycin Concentration (SVC)

A fixed vancomycin loading dose (LD) of 25 mg/kg resulted in a mean initial dose of 1568.25 mg (SD 303.6), with a mean patient weight of 67.4 kg (SD 14.6). The overall mean TTFS was 41.2 hours (SD 13.3), with most TDM samples (85.8%) taken between 24 to 48 hours post-dose.

From 106 TDM samples, only 32.1% achieved the targeted SVC (15.0–20.0 mcg/mL), while 56.6% had subtherapeutic levels (<15.0 mcg/mL) and 11.3% had supratherapeutic levels (>20.0 mcg/mL) (Table 2).

**Table 2:**
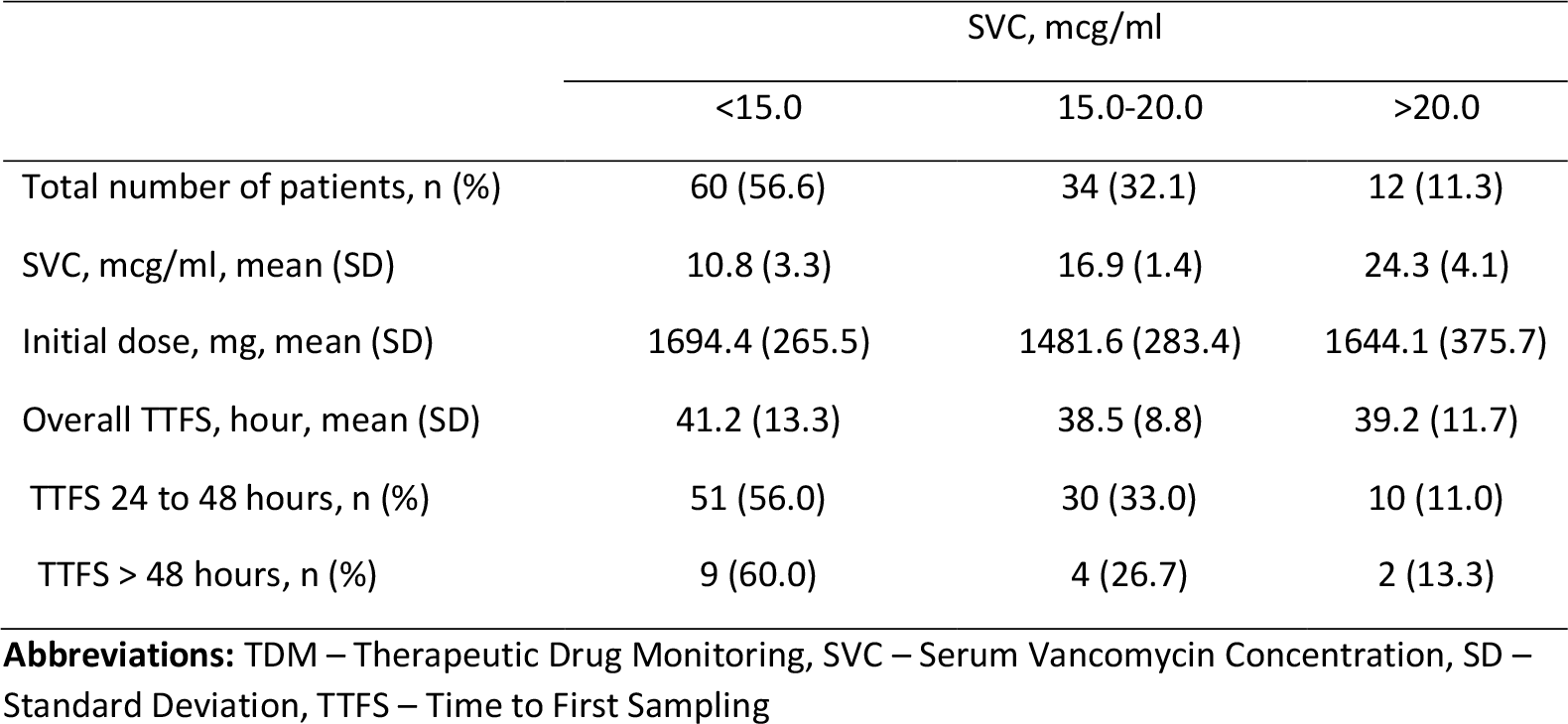
Vancomycin Dosing According to SVC.

|  | SVC, mcg/ml |  |  |
| --- | --- | --- | --- |
|  | <15.0 | 15.0-20.0 | >20.0 |
| Total number of patients, n (%) | 60 (56.6) | 34 (32.1) | 12 (11.3) |
| SVC, mcg/ml, mean (SD) | 10.8 (3.3) | 16.9 (1.4) | 24.3 (4.1) |
| Initial dose, mg, mean (SD) | 1694.4 (265.5) | 1481.6 (283.4) | 1644.1 (375.7) |
| Overall TTFS, hour, mean (SD) | 41.2 (13.3) | 38.5 (8.8) | 39.2 (11.7) |
| TTFS 24 to 48 hours, n (%) | 51 (56.0) | 30 (33.0) | 10 (11.0) |
| TTFS > 48 hours, n (%) | 9 (60.0) | 4 (26.7) | 2 (13.3) |

**Abbreviations:** TDM – Therapeutic Drug Monitoring, SVC – Serum Vancomycin Concentration, SD – Standard Deviation, TTFS – Time to First Sampling

With a fixed LD of 25 mg/kg, the mean SVC in the target range was 16.9 mcg/mL (SD 1.4), while the mean SVC in the supratherapeutic group was 24.3 mcg/mL (SD 4.1). The mean TTFS was comparable across subtherapeutic, target, and supratherapeutic groups. However, more than half (56%) of samples taken between 24 to 48 hours post-dose exhibited subtherapeutic SVC.

Table 3 showed factors that significantly influenced the SVC following the administration of vancomycin initial dose. Simple linear regression was conducted to see the individual effect of each factor on the SVC. The result showed that only age and TTFS had a statistically significant effect on SVC while initial dose showed a slight positive effect. The three factors were then included in the multiple linear regression and all of them able to show a significant effect on SVC; TTFS (p=0.003), age (p= 0.002) and initial dose of vancomycin (p= 0.004) where the association showed in model SVC= 18.809 - 0.132 (TTFS) - 0.125 (Age) + 0.005(Dose)

**Table 3:**
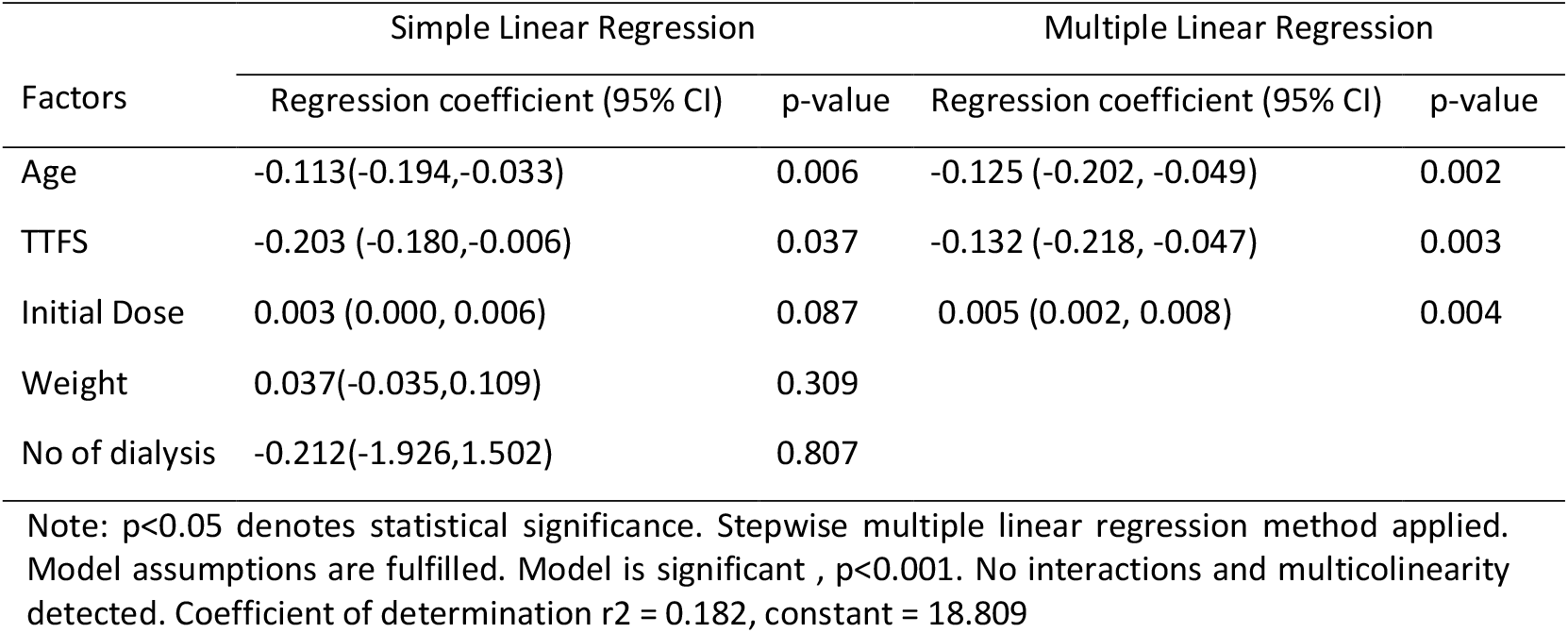
Factors affecting the SVC following initial dose of vancomycin among ESRF patients on dialysis.

## Discussion

The results of this study suggest that the current vancomycin dosing protocol at HPSF does not consistently achieve therapeutic SVC in HD patients with CRBSI. The finding that only 32.1% of patients reached the target SVC range highlights the need for a dosing adjustment. The high proportion (56.6%) of patients with subtherapeutic SVC indicates that the standard LD of 25 mg/kg may be insufficient for most patients, leading to inadequate bacterial eradication and potential treatment failure.

This issue has been similarly reported in previous studies. For example, Rybak et al. found that standard vancomycin dosing in HD patients often resulted in subtherapeutic SVC levels, recommending an increased LD of 20-35 mg/kg to improve outcomes [8]. Additionally, a study by Rosini et. al reported that a 30 mg/kg loading dose of vancomycin resulted in a higher percentage of therapeutic levels at 12 hours compared to the standard 15 mg/kg dose, without an increase in nephrotoxicity or adverse events [11]. Achieving therapeutic drug concentrations rapidly at the early treatment is essential for optimizing clinical outcomes, as it helps reduce the risk of treatment failure and prevents the persistence of bacteria, which could lead to prolonged infection and potential complications [14]. Abdul Gafor et al. further highlighted that inadequate vancomycin exposure was associated with prolonged infection resolution times, reinforcing the need for individualized dosing strategies [6].

Although 11.3% of patients had supratherapeutic levels, this is of less concern in the hemodialysis setting. Vancomycin is typically administered post-dialysis, and approximately 30% of the drug is removed during a single dialysis session. A study by Ariano et al. demonstrated that supratherapeutic pre-dialysis vancomycin levels were often mitigated by dialysis, preventing significant toxicity while maintaining therapeutic efficacy [12]. Similarly, Petejova N. et al. reported that vancomycin clearance during low-flux hemodialysis remove considerable amounts of vancomycin in critically ill septic patients with AKI which reduces toxicity risks, supporting the argument that supratherapeutic levels do not necessarily translate to increased adverse effects in dialysis patients [13]. Therefore, slight elevations in pre-dialysis vancomycin levels should not be overemphasized, as post-dialysis drug removal often brings levels back into the therapeutic range.

The mean initial dose of 1568.2 mg (25 mg/kg) was insufficient for many patients, particularly older individuals and those with delayed TDM sampling. The regression analysis showed a positive association between initial dose and SVC levels, suggesting that increasing the LD to 25-35 mg/kg may improve therapeutic outcomes. This is supported by recent guidelines recommending higher initial doses in HD patients to account for reduced drug clearance [6].

A critical factor influencing SVC was TTFS, which had a significant negative correlation (p=0.003) with SVC levels. Our study found that an optimal TTFS within 24-48 hours post-administration is necessary to accurately assess vancomycin levels and guide subsequent dosing. Delayed sampling beyond 48 hours was associated with greater fluctuations in SVC, which may lead to inappropriate dose modifications.

Age also played a significant role in vancomycin pharmacokinetics, with older patients exhibiting lower SVCs (p=0.002). This finding aligns with previous studies that suggest age-related changes in renal function and altered drug distribution can impact vancomycin clearance, necessitating dose adjustments in elderly patients. Furthermore, the initial vancomycin dose demonstrated a mild positive correlation with SVC (p=0.004), indicating that higher loading doses could improve target attainment.

Compared with prior research, our study supports the recommendations of Rybak et al., which suggested that an increased LD of 20-35 mg/kg may be necessary to achieve target vancomycin levels in HD patients [8]. Similarly, Mermel et al. emphasized the importance of precise TDM sampling, reinforcing our conclusion that optimizing TTFS is crucial for therapeutic success [7].

The variability in SVC observed in our study suggests that a fixed dosing strategy may not be ideal for all patients. Instead, individualized dosing models incorporating patient-specific pharmacokinetic parameters, weight, and dialysis efficiency may yield better therapeutic outcomes. Implementing a revised protocol with an increased LD (25-35 mg/kg) and standardized TTFS within 24-48 hours post-dose could enhance the achievement of target SVC and reduce the risk of treatment failure or toxicity.

## Conclusion

The current vancomycin dosing protocol at HPSF is suboptimal in achieving therapeutic SVC in HD patients with CRBSI. An increase in the LD to 25-35 mg/kg and optimized TTFS may enhance vancomycin efficacy. Further prospective studies are needed to validate these findings and refine dosing recommendations for improved clinical outcomes.

## Data Availability

All data produced in the present study are available upon reasonable request to the authors

## Acknowledgement

We would like to thank the Director General of Health, Malaysia, for his permission to publish the findings from this study. We also would like to thank the head and staff of the Therapeutic Drug Monitoring Unit, Pharmacy Department of the Hospital Pakar Sultanah Fatimah for the permission and assisting us in the data collection

## Notes

### Competing Interest Statement

The authors have declared no competing interest.

### Funding Statement

This study did not receive any funding

### Author Declarations

Ethics committee/IRB of Medical Research Ethics Committee Malaysia gave ethical approval for this work with reference number 24-02412-PBR

## References

1. Eknoyan, G., et al., The burden of kidney disease: improving global outcomes. Kidney Int, 2004. 66(4): p. 1310–4.

2. 18th Report of the Malaysian Dialysis and Transplant Registry 2010 â“NRR â” National Renal Registry. 2011; Available from: https://www.msn.org.my/nrr/18th-report-of-the-malaysian-dialysis-and-transplant-registry-2010/.

3. 22nd Report of the Malaysian Dialysis and Transplant Registry 2014 â“NRR â” National Renal Registry. 2015; Available from: https://www.msn.org.my/nrr/22nd-report-of-the-malaysian-dialysis-and-transplant-registry-2014/.

4. Shingarev, R., J. Barker-Finkel, and M. Allon, Natural history of tunneled dialysis catheters placed for hemodialysis initiation. J Vasc Interv Radiol, 2013. 24(9): p. 1289–94.

5. Allon, M., Dialysis catheter-related bacteremia: treatment and prophylaxis. Am J Kidney Dis, 2004. 44(5): p. 779–91.

6. Abdul Gafor, A.H., et al., Antibiogram for haemodialysis catheter-related bloodstream infections. Int J Nephrol, 2014. 2014: p. 629459.

7. Mermel, L.A., et al., Clinical practice guidelines for the diagnosis and management of intravascular catheter-related infection: 2009 Update by the Infectious Diseases Society of America. Clin Infect Dis, 2009. 49(1): p. 1–45.

8. Rybak, M.J., et al., Therapeutic monitoring of vancomycin for serious methicillin-resistant Staphylococcus aureus infections: A revised consensus guideline and review by the American Society of Health-System Pharmacists, the Infectious Diseases Society of America, the Pediatric Infectious Diseases Society, and the Society of Infectious Diseases Pharmacists. Am J Health Syst Pharm, 2020. 77(11): p. 835–864.

9. Oshvandi, K., et al., High-flux and low-flux membranes: efficacy in hemodialysis. Nurs Midwifery Stud, 2014. 3(3): p. e21764.

10. Abe, M., et al., Super high-flux membrane dialyzers improve mortality in patients on hemodialysis: a 3-year nationwide cohort study. Clin Kidney J, 2022. 15(3): p. 473–483.

11. Rosini, J.M., et al., A randomized trial of loading vancomycin in the emergency department. Ann Pharmacother, 2015. 49(1): p. 6–13.

12. Ariano, R.E., et al., Adequacy of a vancomycin dosing regimen in patients receiving high-flux hemodialysis. Am J Kidney Dis, 2005. 46(4): p. 681–7.

13. Petejova, N., et al., Vancomycin removal during low-flux and high-flux extended daily hemodialysis in critically ill septic patients. Biomed Pap Med Fac Univ Palacky Olomouc Czech Repub, 2012. 156(4): p. 342–7.

14. Bruniera, F.R., et al., The use of vancomycin with its therapeutic and adverse effects: a review. Eur Rev Med Pharmacol Sci, 2015. 19(4): p. 694–700.

